# A dia-PASEF workflow improves proteome coverage and quantitative performance in repeated-measures analysis of cervicovaginal fluid from a preconception cohort (EARLY-PREG)

**DOI:** 10.1101/2025.05.19.25327800

**Authors:** Mauricio Hernandez, Pablo Saldivia, Veronica Latapiat, Barbara Antilef, Guillermo Nourdin, Felipe Castro, Cristian Vargas, Elard S. Koch

**Author notes:** These authors contributed equally to this work (Mauricio Hernandez, Pablo Saldivia, Veronica Latapiat). **Corresponding author:** Elard S. Koch, MELISA Institute, Dalcahue 1120, San Pedro de la Paz, 4133515, Concepción, Chile.

## Abstract

Cervicovaginal fluid (CVF) represents a valuable source of molecular information reflecting time-dependent proteomic changes across the menstrual cycle. However, the large-scale discovery of CVF biomarkers remains challenging because of limitations in the sensitivity, reproducibility, and data completeness of mass spectrometry-based workflows, particularly within repeated-measures longitudinal study designs. Here, we evaluated a standardised data-independent acquisition (DIA)-based proteomics workflow (dia-PASEF) for longitudinal proteomic analysis of serial CVF samples collected across an ovulation-anchored seven-day window (d0-d6) from a prospective preconception cohort (EARLY-PREG). The performance of this dia-PASEF approach was compared with that of a dda-FracOffline strategy. Across the time window, 2,817 quantifiable proteins were detected with dda-FracOffline, whereas 4,229 quantifiable proteins were detected with dia-PASEF, with mean geometric coefficient of variation values of 203.1% and 55.6%, respectively. Using day 0 (ovulation) as a reference for day-to-day comparisons, more differentially expressed proteins (DEPs) were identified with dia-PASEF than with dda-FracOffline across pairwise comparisons (952–1,116 DEPs for dia-PASEF versus 161–460 DEPs for dda-FracOffline). Pathway analyses demonstrated broader biological coverage with dia-PASEF. In conclusion, dia-PASEF is a sensitive and reproducible approach for longitudinal repeated-measures proteomic analysis of CVF. This method is adaptable to other biofluids and supports the investigation of time-dependent molecular signatures and associated biological pathways under physiologically relevant conditions in female reproductive system research.

## Introduction

Proteomic analysis of female reproductive tissues is being increasingly recognised as a relevant approach for studying women’s fertility and reproductive health. At the molecular level, insights gained from proteomic studies of tissues and biofluids^1^ may contribute to our understanding of pathophysiological processes affecting different biological systems. Despite its importance, the proteome and its time-dependent molecular changes in female reproductive fluids and tissues under physiological and pathological conditions remain underexplored ^2–4^. Furthermore, despite the accelerated development of proteomics research, the discovery of reliable biomarkers remains challenging, due to methodological and analytical limitations affecting longitudinal repeated-measures designs ^5^.

Cervicovaginal fluid (CVF) is recognised as a relevant biofluid source for biomarker discovery in proteomics research because of its continuity with the female reproductive tract and pivotal role in fertility, pregnancy outcomes, and local immune responses ^6–11,12,13^. As a non-invasive source of biological information, CVF reflects physiological and pathological changes, including the menstrual cycle, pregnancy, and disease progression. Recent studies have linked specific proteins in CVF to endometriosis ^14,15^, pregnancy-related complications ^16,17^ and several cancer types, such as ovarian, endometrial, and cervical malignancies ^6–11^.

Recent advances in proteomics techniques have advanced the identification of protein and quantification of their levels. However, most studies rely on data-dependent acquisition (DDA), a time-consuming workflow with limitations in quantitative capabilities, reproducibility, and accuracy, particularly in large-scale longitudinal analyses with repeated measures over time^18^. In the search for new strategies for more extensive proteome coverage, data-independent acquisition (DIA)-based methodologies have recently been developed^19^. For example, Njoku and collaborators ^7^ recently applied a DIA method using sequential window acquisition of all theoretical fragment ion mass spectra (SWATH-MS) to analyse CVF samples, which enabled the quantification of the levels of 2,425 proteins, a greater number of proteins than could be quantitatively analysed via a standard DDA approach.

Addressing the limitations of DDA methods is important for advancing the epidemiological and clinical applications of CVF proteomic analysis, particularly through longitudinal cohort studies ^20,21^ that enable the identification of biomarkers and the characterisation of time-dependent pathophysiological changes ^22,23^. In fact, following biomarker variation over time may reveal molecular signatures linked to specific biological processes, offering more comprehensive insights into female reproductive health and disease. Despite the potential of DIA, studies involving the use of spectral library-based DIA to analyse CVF are rare compared with studies involving the use of traditional DDA, which has long been the dominant approach ^24,25^. Furthermore, to date, no proteomic studies have systematically evaluated longitudinal changes in the CVF proteome across consecutive days of the human menstrual cycle.

Although SWATH-MS has been explored as an alternative approach to DIA to assess the CVF proteome, its reliance on fixed isolation windows may introduce spectral interference that potentially limits resolution. With respect to new technologies that address this limitation, in 4D proteomics, enabled by timsTOF mass spectrometry (MS), trapped ion mobility spectrometry (TIMS) is integrated with parallel accumulation–serial fragmentation (PASEF), increasing proteome resolution and coverage ^26–28^. In this context, DIA strategies such as parallel accumulation–serial fragmentation combined with data-independent acquisition (dia-PASEF) have been applied as quantitative research tools ^29^ for complex biofluids such as plasma; in this strategy, ion mobility enables improved separation of ions on the basis of their shape, size, and charge ^26^.

In this study, we aimed to implement and validate an optimised and reproducible dia-PASEF method for time-dependent proteomic analyses of serial CVF samples collected throughout the menstrual cycles of women participating in the EARLY-PREG open longitudinal preconception cohort (NCT07358026) ^30^. The dia-PASEF approach was compared with the traditional DDA approach coupled with offline peptide fractionation (dda-FracOffline). We hypothesised that the dia-PASEF approach would provide greater proteome coverage and reproducibility than traditional DDA, supporting its potential application for biomarker discovery in repeated-measures longitudinal studies of women’s fertility, reproductive health, and pregnancy.

## Methods

### 1. EARLY-PREG cohort biobank

EARLY-PREG (ClinicalTrials.gov identifier NCT07358026) is a preconception longitudinal open cohort designed to investigate female menstrual cycles, fertility and maternal–embryonic communication at the early stages of pregnancy during the peri-implantation window. The EARLY-PREG biobank is a longitudinal biobank that constitutes a structured and temporally indexed repository of multiple maternal biological samples, including CVF, urine, blood fractions, and saliva. In this MS validation study, synchronised and well-documented CVF samples from the EARLY-PREG biobank were used to compare dia-PASEF with the traditional DDA approach coupled with offline peptide fractionation (dda-FracOffline).

### 2. Longitudinal CVF samples

At present, the EARLY-PREG biobank includes different biofluids from female volunteers (n = 223) collected daily during their menstrual cycles from the end of menstruation, which represents the beginning of a new cycle. Ovulation was determined by quantifying the surge in luteinising hormone (LH) levels in urine. Samples were collected from the day after the end of menstruation until the 14th day post-ovulation, as determined by the surge in LH levels during each cycle, implementing a previously described method ^31^ used to detect ovulation within ±1 day (95.7% confidence). LH concentrations were determined through a radioimmunoassay (IRMA) (Barnafi-Krause Laboratory, Santiago, Chile). Through this approach, the day of the surge in LH levels was defined as day 0 (d0), which served as a cycle-anchored time reference for synchronising repeated measures across individuals.

### 3. Sample size

This study was designed as a methodological validation of the dia-PASEF workflow to assess its ability to capture longitudinal proteomic changes in cervicovaginal fluid (CVF) during the human menstrual cycle, using offline-fractionated DDA-MS as the reference analytical approach rather than as a population-based biomarker discovery study. Consequently, the primary objective was to maximise internal validity and analytical comparability while minimising biological variability. First, to enable within-individual repeated-measures analyses, we predefined an ovulation-anchored luteal phase window comprising seven consecutive days (d0–d6). Participants were selected exclusively based on the availability of CVF samples for all days across this period. Women with a luteal phase shorter than seven days or for whom stored CVF samples from the selected window were missing were excluded. Second, to further reduce biological heterogeneity and heteroscedasticity in longitudinal analyses, participants were selected according to strict matching criteria, including age, body mass index, menstrual cycle length, bleeding duration, completeness of longitudinal sampling, and other relevant reproductive characteristics. Finally, only women with samples from non-conception cycles available in the EARLY-PREG biobank were selected to avoid pregnancy-related physiological changes during the analysed window; the final analysed cohort comprised six participants (42 longitudinal CVF samples). The characteristics of the selected participants are summarised in Table 1.

**Table 1:** Characteristics of the women whose samples were used in the proteomic experiments.

| Factors | Women (n = 6) |  |  |  |  |  |
| --- | --- | --- | --- | --- | --- | --- |
|  | P1 | P2 | P3 | P4 | P5 | P6 |
| Age (mean ± SD) | 32.3 ± 3.6 |  |  |  |  |  |
| Weight (kg) | 62 | 57 | 66 | 67 | 60 | 74 |
| Height (m) | 1.52 | 1.60 | 1.68 | 1.61 | 1.58 | 1.7 |
| BMI<sup>1</sup> | 26.8 | 22.2 | 24.4 | 25.8 | 24 | 25.6 |
| Waist circumference (cm) | 72 | 64 | 66 | 70 | 68 | 82 |
| Age at menarche (mean ± SD) | 12.7 ± 1.2 |  |  |  |  |  |
| Menstrual cycle length<sup>2</sup> (days) | 28 | 30 | 24 | 30 | 29 | 33 |
| Menstruation length<sup>3</sup> (days) | 5 | 6 | 6 | 6 | 6 | 6 |

<sup>1</sup>Body mass index (BMI) was calculated as weight (kg) divided by height squared (m<sup>2</sup>). <sup>2</sup>Menstrual cycle length refers to the number of days from the first day of menstruation to the first day of the next cycle. <sup>3</sup>Menstruation duration corresponds to the number of days of menstrual bleeding per cycle.

### 4. Protein extraction and processing for MS analysis

CVF aliquots (100–500 µL) were thawed on ice and combined with equal volumes of 100 mM NH4HCO3 and 100 mM DTT. The mixtures were homogenised by vortexing, incubated at 95 °C for 15 minutes, and cooled on ice for 5 minutes. The samples were then subjected to cold ultrasound disruption at 4 °C (10-second on/off pulses at 50% amplitude) and subsequently centrifuged at 19,000 × g for 20 minutes at 4 °C. The supernatant (soluble phase) was carefully separated, while the pellet (insoluble phase) was retained for further processing.

The soluble phase was concentrated in Vivaspin tubes (10,000 MWCO filter, Sartorius), washed with 400 µL of ultrapure water, and centrifuged at 12,000 × g for 10 minutes at 4 °C. The filtrate (<10,000 Da) was dried in a SpeedVac and stored at −20 °C, while proteins retained on the filter (>10,000 Da) were recovered by washing with ∼600 µL of water, combined, and dried. The insoluble pellet and soluble protein concentrate were resuspended in 8 M urea with 25 mM NH₄HCO₃. The mixture was sonicated on ice and centrifuged at 19,000 × g for 15 minutes at 4 °C. The protein content was quantified using a Qubit protein assay (Invitrogen, #Q33212).

For MS preparation, 300 µg of total protein from the soluble and insoluble phases was reduced with 20 mM DTT in 25 mM NH₄HCO₃ at 25 °C for 1 hour. Alkylation was performed with 20 mM iodoacetamide in 25 mM NH₄HCO₃ in the dark for 1 hour. The samples were diluted eightfold with 25 mM NH₄HCO₃ and digested with sequencing-grade trypsin (#V5071; Promega) at a 1:50 protease/protein ratio for 16 hours at 37 °C. Digestion was stopped by the addition of 10% formic acid. Peptides were cleaned using Sep-Pak C18 Spin columns (Waters), dried in a rotary concentrator at 40 °C, and stored for subsequent MS analysis.

### 5. dda-FracOffline

#### 5.1. Peptide Fractionation

Offline high-pH reversed-phase fractionation of CVF peptides was performed using a protocol adapted from our previously published tear proteomics workflow ^32^. A total of 200 µg of tryptic peptides from soluble and insoluble CVF phases were separated using an ÄKTA avant 25 system, with refrigerated fraction collection at 6 °C. Peptides were loaded onto a BEH reversed-phase column (2.1 mm × 5 cm, Waters) at a pH of 10 and a flow rate of 0.2 mL/min. Buffer A consisted of 5 mM ammonium formate in water, whereas buffer B consisted of 5 mM ammonium formate in 90% ACN. A binary gradient in which the buffer B concentration increased linearly from 3% to 40% over 30 minutes and then to 60% over 15 minutes and to 85% in the final 5 minutes was applied. We collected a total of 24 fractions (400 µL each), which were vacuum-dried and then reconstituted in water containing 2% ACN and 0.1% FA. These fractions were concatenated into 8 final fractions.

#### 5.2. Mass Spectrometry

nLC–MS analysis was performed on a nanoELUTE (Bruker Daltonics) system coupled online with a hybrid TIMS-quadrupole TOF mass spectrometer, timsTOF Pro (Bruker Daltonics, Germany), using a nanoelectrospray ion source (CaptiveSpray, Bruker Daltonics) ^33^. For long-gradient nLC–MS/MS runs, we followed the standard protocol described by Meier et al., 2021. Approximately 200 ng of peptides were separated on an Aurora series reversed-phase column (25 cm × 75 µm ID, 1.9 µm; IonOpticks, Australia) at a flow rate of 400 nL/min in an oven compartment heated to 50 °C. For the analysis of off-line fractionated CVF samples, we applied a multistep gradient beginning with an increase in the buffer B concentration from 2% to 17% over 60 minutes, to 25% in 30 minutes, to 37% in 10 minutes, and finally to 95% in 10 min (held for 10 min). The column was equilibrated using 4 volumes of solvent A. The mass spectrometer was operated in data-dependent PASEF ^34^ mode, including 1 TIMS-MS survey and 10 PASEF MS/MS scans per acquisition cycle. Mobility values were acquired across a 1/K0 range of 1.6 to 0.6 Vs cm-2, using equal ion accumulation and a ramp time of 100 ms each for the dual-TIMS analyser.

#### 5.3. Data searching

Computational processing of the DDA data was performed using FragPipe v19.1 software. The pipeline integrates the MSFragger (version 3.7)^35^ search engine for peptide-spectrum matching, Philosopher (version 3.4.13)^36^ for False Discovery Rate (FDR) control and validation, and EasyPQP (https://github.com/grosenberger/easypqp; version 0.1.9) for spectral library generation. Raw (.d) files were used as inputs for MSFragger searches. Protein identification was conducted against the reference proteome of *Homo sapiens* (UP000005640) from UniProt/SwissProt (reviewed sequences only; downloaded on Feb. 15, 2021), supplemented with a database of common contaminant proteins in MS experiments available in FragPipe. Decoy sequences were generated using reversed protein sequence entries to enable target-decoy estimation. For MSFragger analysis, the mass tolerance for precursor and fragment ions was set to 20 ppm. The enzyme specificity was defined as ‘strict trypsin’, allowing full tryptic cleavage and a maximum of two missed cleavages. Carbamidomethylation of cysteine was included as a fixed modification, whereas methionine oxidation and N-terminal acetylation were set as variable modifications. The final spectral library was filtered at 1% FDR at both the peptide and protein levels.

### 6. dia-PASEF

#### 6.1. Mass spectrometry

The timsTOF Pro mass spectrometer was operated in dia-PASEF mode with the following settings: mass range, 100 to 1700 m/z; ion mobility range, 1/K0 start, 0.57 V/cm2; end, 1.47 V/cm2; ramp and accumulation times, 100 ms; capillary voltage, 1600 V; dry gas, 3 L/min; and dry temperature, 160 °C. The dia-PASEF ion mobility parameters were as follows: Each cycle consisted of one MS1 full scan and 64 MS2 windows covering 400–1200 m/z and 0.57–1.46 1/K0. Each window was 25 Th wide by 0.2 V s/cm2 high. There was an overlap of 0.05 in 1/K0 and no overlap in m/z. The cycle time was 1.80 seconds. The CID collision energy ranged from 20 to 52 eV as a function of the inverse mobility of the precursor ^24^. Each sample was injected into a nanoELUTE nanoliquid chromatography system (Bruker Daltonics). Peptides (200 ng of digest) were separated within 60 min at a flow rate of 400 nL/min on an Aurora Series CSI reversed-phase column (25 cm × 75 µm i.d., C18, 1.6 µm; IonOpticks, Australia) at 50 °C. Mobile phases A and B were water and acetonitrile with 0.1 vol% FA, respectively. The B concentration was linearly increased from 2 to 17% within 37 min, followed by an increase to 25% within 15 min and further to 35% within 8 min, followed by a washing step at 85% B and re-equilibration.

#### 6.2. Data search

To construct the DDA spectral library, a pool of tryptic peptides from all the samples was used. For peptide fractionation, mass spectrometry and data searching, the methods described in the DDA section were used (see Methods sections 5.1, 5.2 and 5.3). Proteomic data analyses were performed on a high-performance computing server with dual AMD EPYC 7552 48-core processors (2.20 GHz) and 512 GB of RAM. Raw dia-PASEF data were processed via DIA-NN v1.7.15 software ^37^ using the spectral library generated in the previous step. Mass accuracy thresholds were set to a default average of 10 ppm for MS1 and MS2 spectra. Match-between-runs (MBR) was enabled; the quantification mode was set to “any LC (high accuracy)”. The default settings were used for all remaining parameters. Identified proteins were filtered using a precursor-level q value < 1% and a global protein-level q value < 1%.

### 7. Bioinformatics and statistical analysis

The intensity matrix reports from MSFragger and DIA-NN were exported and processed in the R statistical environment. Proteins were retained when intensity values were present in at least 50% of the samples per analysed category, and missing values were imputed using the missForest algorithm ^38,39^. The imputed matrices were normalised using quantile normalisation and log_2_-transformed. Absolute and relative numbers (%) are reported for longitudinally differentially expressed proteins (DEPs) along the menstrual cycle, and significant differences were determined using Bayesian t tests for pairwise comparisons between days (e.g., d0 vs. dn, d1 vs. dn), considering the normalised intensity means of biological replicates ^40^. Proteins were considered differently expressed at p < 0.05. Exploratory analysis and data visualisation were performed in R (v.3.6.0) using EnhancedVolcano (v.1.2.0), ComplexHeatmap (v.2.0.0), and standard base packages. Because the primary comparisons were performed within individuals across time, each participant served as her own control, reducing between-subject variability and increasing sensitivity to detect temporal proteomic changes. The geometric coefficient of variation (gCV) was calculated across samples for each quantifiable protein, defined as a protein that was consistently detected across each condition and presented measurable relative abundance values. Principal component analysis (PCA) of the intensity matrices was performed. For each sample, the total signal intensity was calculated as the sum of the protein intensities and log_10_-transformed. Pearson correlations between log_10_-transformed total intensity values and PC1 and PC2 scores were calculated. The relative contributions of total signal intensity, participant, and sampling day were estimated using multiple linear regression. Total signal intensity was used as a sample-level technical proxy and was not considered an independent measure of instrument performance.

### 8. Pathway and tissue enrichment analysis

Pathway enrichment analysis was performed using ReactomePA (v.1.42.0), for which the human Reactome database is queried with log2-fold change (logFC) and p value data obtained for each statistical comparison. For the analysis of enrichment across human systems, tissues, and cells, we used the TISSUES 2.0 database ^41^ and the same statistical parameters used for pathway analysis through topGO (v.2.5.0). Boxplots were generated to show the distribution of protein intensities using ggpubr (v.0.6.0). Statistical comparisons between groups were conducted using t tests and chi-square tests with rstatix (v.0.7.2). Adjusted p values obtained from the Reactome enrichment analysis were used to calculate the enrichment score (−log_10_ FDR). The z score was calculated on the basis of the logFC of each protein belonging to the designated reactome and tissue categories using the GOplot (v.1.0.2) package. Importantly, this z score is not equivalent to the conventional z score; instead, it provides a directional estimate indicating whether the proteins associated with a biological process, molecular function, or cellular component tended to be upregulated (positive value) or downregulated (negative value). The z score was determined on the basis of the number of proteins assigned as upregulated (logFC > 0) and those classified as downregulated (logFC < 0), labelled “up” and “down”, respectively, in the dataset.

### 9. Panel of selected biomarkers

To assess the efficiency of dia-PASEF in biomarker detection, an FDA-proposed biomarker panel derived from a previously published study ^42^ was used to directly compare the ability of dia-PASEF and dda-FracOffline to detect these clinically relevant proteins and quantify their abundance. In addition, we conducted an extensive literature-based curation of candidate proteins associated with physiological and pathological conditions affecting the female reproductive tract. Searches were performed using keywords such as “menstrual cycle”, “ovarian carcinoma”, “endometrial carcinoma”, “cervical carcinoma”, and “endometriosis”, integrating evidence from proteomic, transcriptomic, and genomic studies.

### 10. Data availability

The proteomic data can be accessed through the ProteomeXchange Consortium via PRIDE, with the dataset identifiers PXD071350 and PXD071584. The R scripts and analysis workflow are publicly available at GitHub at the following address: https://github.com/MELISAInstitute2025/cvf-proteomics-reproducibility.

### 11. Ethics approval and consent to participate

The EARLY-PREG protocol was approved by the Ethics Committee of the Servicio de Salud de Concepción, Chile (CEC: 17-03-06; NCT Number: NCT07358026). All participants provided written informed consent prior to enrolment. All procedures were conducted in accordance with the Declaration of Helsinki and Good Clinical Practice guidelines.

## RESULTS

### 1. Characterisation of participants and sampling

The detailed characteristics of the participants are provided in Table 1. The length of the menstruation phase of the cycle (5.8 ± 0.4), the total length of the menstrual cycle (29.0 ± 3.0), age (32.3 ± 3.6), and other variables were within comparable ranges (Figure 1A). Forty-two CVF samples (n = 42) stored at −80 °C were processed for protein extraction and tryptic digestion and then analysed using the two proteomic workflows (Figure 1B) as described in the Methods section. All 42 samples were analysed on a timsTOF Pro mass spectrometer over a two-week acquisition period. This included 336 chromatographic runs using dda-FracOffline and 42 direct runs using dia-PASEF. For each analytical workflow, this design yielded a within-individual repeated-measures time series comprising a 7-day ovulation-anchored window (d0–d6) of CVF proteomes per participant.

**Figure 1.**
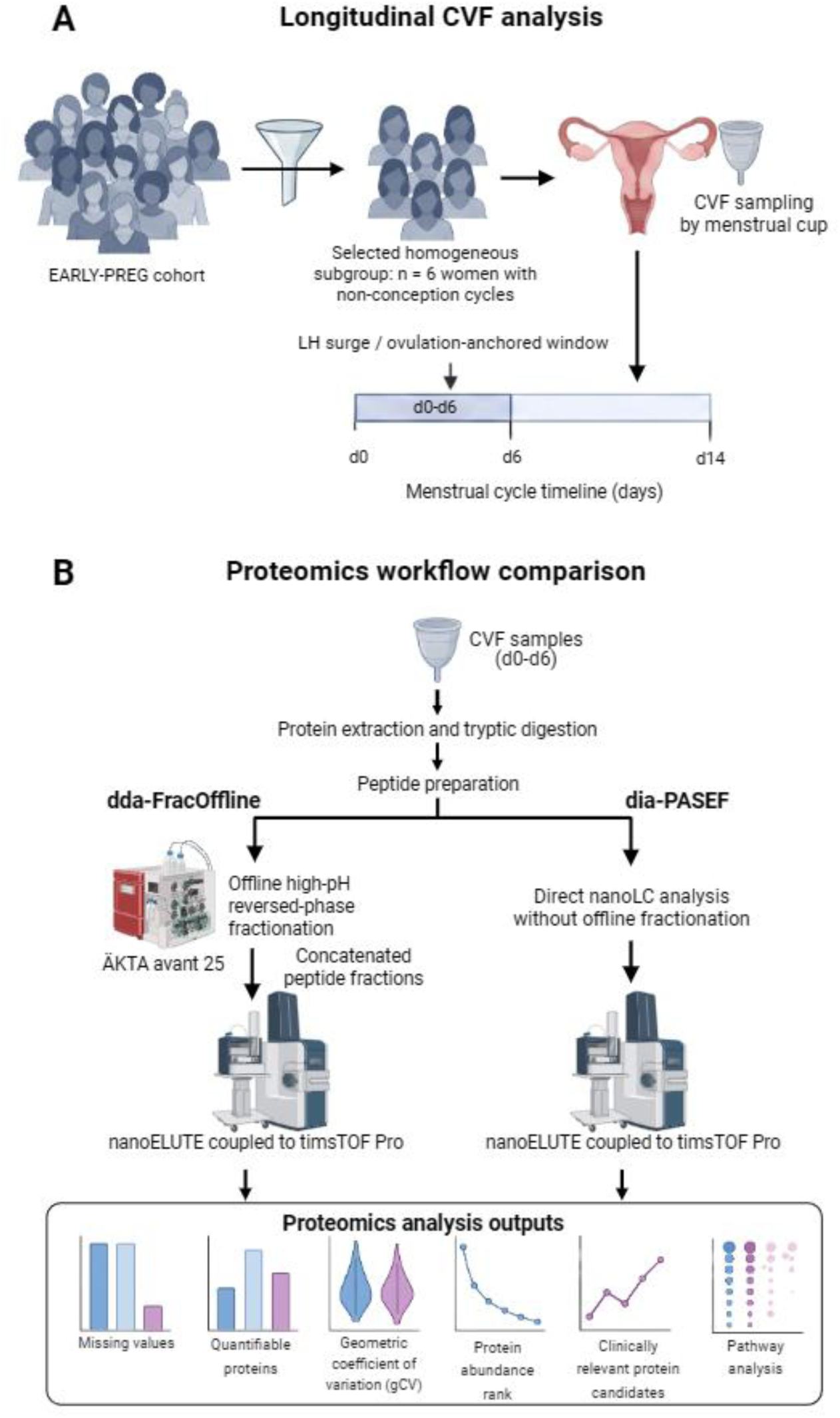
Proteomic workflow for the longitudinal repeated-measures analysis of CVF. Schematic representation of the experimental design and proteomic workflows used to analyse CVF samples from the EARLY-PREG cohort. (A) A homogeneous subgroup of six women with non-conception cycles was selected, and CVF samples collected across a 7-day ovulation-anchored window (d0–d6) were analysed using a within-individual repeated-measures design. (B) Following protein extraction and tryptic digestion, peptide preparations were analysed using dda-FracOffline or dia-PASEF. dda-FracOffline included offline high-pH reversed-phase fractionation, whereas dia-PASEF involved direct nanoLC–MS/MS analysis without offline fractionation. The workflows were compared according to missing values, quantifiable proteins, geometric coefficient of variation (gCV), protein abundance rank, clinically relevant protein candidates, and pathway analysis. Hernandez, M. (2026). Created in BioRender. https://BioRender.com/ifbud0f

### 2. Quantitative characteristics of the dda-FracOffline and dia-PASEF datasets

We conducted a comparative analysis focusing on dataset-level quantitative characteristics, including intersample correlations, the proportion of missing values, and protein identification and quantification performance. First, pairwise correlations between biological samples were assessed as a descriptive measure of dataset consistency. The mean correlation coefficient was approximately 0.70 for dda-FracOffline and 0.90 for dia-PASEF (Figure S2). Second, we evaluated the proportion of missing values across samples. The dia-PASEF workflow yielded a lower proportion of missing values than dda-FracOffline, with some dda-FracOffline samples reaching up to 75% missing values (Figure 2A). Third, correlation analysis across samples demonstrated lower intersample consistency for dda-FracOffline than for dia-PASEF (Figure 2B). Next, across the ovulation-anchored time series (d0–d6), between 1,491 and 7,183 proteins across time points were identified by dda-FracOffline (Figure 2C), whereas between 3,811 and 5,398 proteins across time points were detected by dia-PASEF (Figure 2D). Finally, we analysed protein identification and quantification performance. A total of 9,593 proteins were identified by dda-FracOffline, whereas 5,986 proteins were identified by dia-PASEF. However, when considering quantifiable proteins (identified and consistently measured across samples), 2,817 proteins were identified by dda-FracOffline, whereas 4,229 proteins were identified by dia-PASEF (Figure 2E, left), corresponding to a 50% higher quantification yield for dia-PASEF. Overall, both workflows yielded 4,316 unique quantifiable proteins, including 2,730 shared proteins, 87 unique to dda-FracOffline, and 1,499 unique to dia-PASEF (Figure 2E). The gCV for quantifiable proteins—which was calculated across samples for each protein—was lower for dia-PASEF than for dda-FracOffline (mean 55.6% vs. 203.1%, respectively; Figure 2E, centre), indicating lower across-sample quantitative variability in the dia-PASEF dataset. Dataset-level analysis revealed a stronger association between PCA variation and total signal intensity in dda-FracOffline than in dia-PASEF, particularly for PC1 (r = 0.90 vs. 0.24). Consistently, total signal intensity accounted for 29.6% of the modelled variation for dda-FracOffline compared with 6.5% for dia-PASEF, with additional contributions from participant and sampling day (Figure S1).

**Figure 2.**
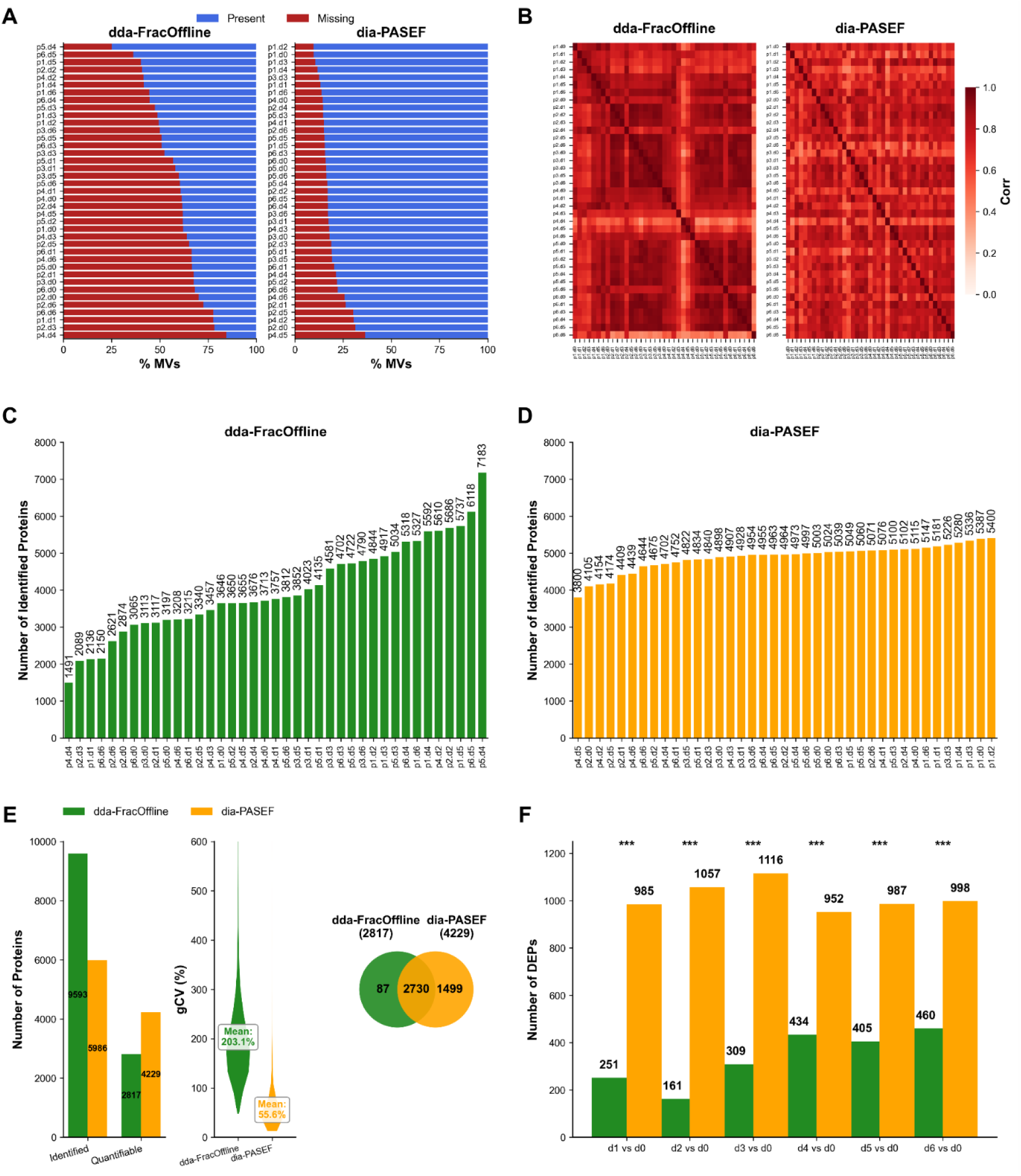
Dataset-level quantitative consistency and evaluation of proteomic data obtained by dda-FracOffline and dia-PASEF. (A) Proportion of missing values (%MVs) for each sample (present, blue; missing, red). (B) Pearson correlation heatmaps between samples (positive correlation, red; no correlation, white). (C, D) Number of identified proteins per sample for dda-FracOffline (C, green) and dia-PASEF (D, orange). (E) Overview of identified and quantifiable proteins (left), geometric coefficient of variation (gCV) distributions across quantifiable proteins (centre), and shared and unique quantifiable proteins (right). (F) Number of differentially expressed proteins (DEPs) identified by dda-FracOffline (green) and dia-PASEF (orange) in longitudinal comparisons against d0 (***, p < 0.001; chi-square test).

Although broader overall protein identification was achieved with dda-FracOffline, dia-PASEF demonstrated greater consistency in protein detection across the ovulation-anchored repeated-measures time series.

#### 2.1. Differential Protein Expression

Following dataset-level quantitative assessment, we assessed the time-dependent changes in differentially expressed proteins (DEPs) across the menstrual cycle to characterise variations in protein abundance in CVF within the ovulation-anchored, repeated-measures 7-day window (*d0–d6*). Comparative analyses revealed significant differences between methodologies in the detection and quantification of DEPs across longitudinal CVF samples (Figure S3). At most time points, compared with dda-FracOffline, dia-PASEF resulted in higher absolute and relative numbers of DEPs, e.g., 985 (23%) vs. 251 (8.9%) between *d0* and *d1* and 1,057 (24.9%) vs. 161 (5.7%) between *d0* and *d2*. From d0 to d6, dia-PASEF maintained a more stable detection rate (Figure 2F). These findings indicate that dia-PASEF enables more consistent detection and quantification of differentially expressed proteins in longitudinal repeated-measures designs, facilitating the capture of time-dependent proteomic changes across the menstrual cycle (Figure 2F and Figure S3).

### 3. Longitudinal enrichment analysis of pathways, systems, tissues, and cells

To interpret the functional relevance of the quantified proteins, we performed tissue and pathway enrichment analyses. We evaluated tissue representation on the basis of DEPs identified across longitudinal comparisons of CVF collected within the ovulation-anchored repeated-measures window (Figure S4). The analysis revealed enrichment in tissues associated with the female reproductive system, including the uterus and uterine cervix, as well as immune-related tissues. The dia-PASEF workflow demonstrated broader and more consistent tissue coverage, particularly at later time points (*d5* and *d6*), with elevated enrichment scores (−log_10_ adjusted p value) and the highest protein densities. We further examined the biological processes associated with the differential protein profiles quantified across the 7-day window. Common pathways identified with both strategies between time points *d0* and *d6* (Figure 3) included ‘Formation of the cornified envelope’, ‘Metabolism of proteins’, and ‘Processing of capped intron-containing pre-mRNA’. Additional pathways, such as ‘Signalling by RHO GTPases’ and ‘Mitotic Metaphase/Anaphase Transition’, were related to cell signalling and cell-cycle-related processes (Figure 3). Compared with dda-FracOffline, dia-PASEF showed increased pathway coverage and enrichment across the time-dependent series.

**Figure 3.**
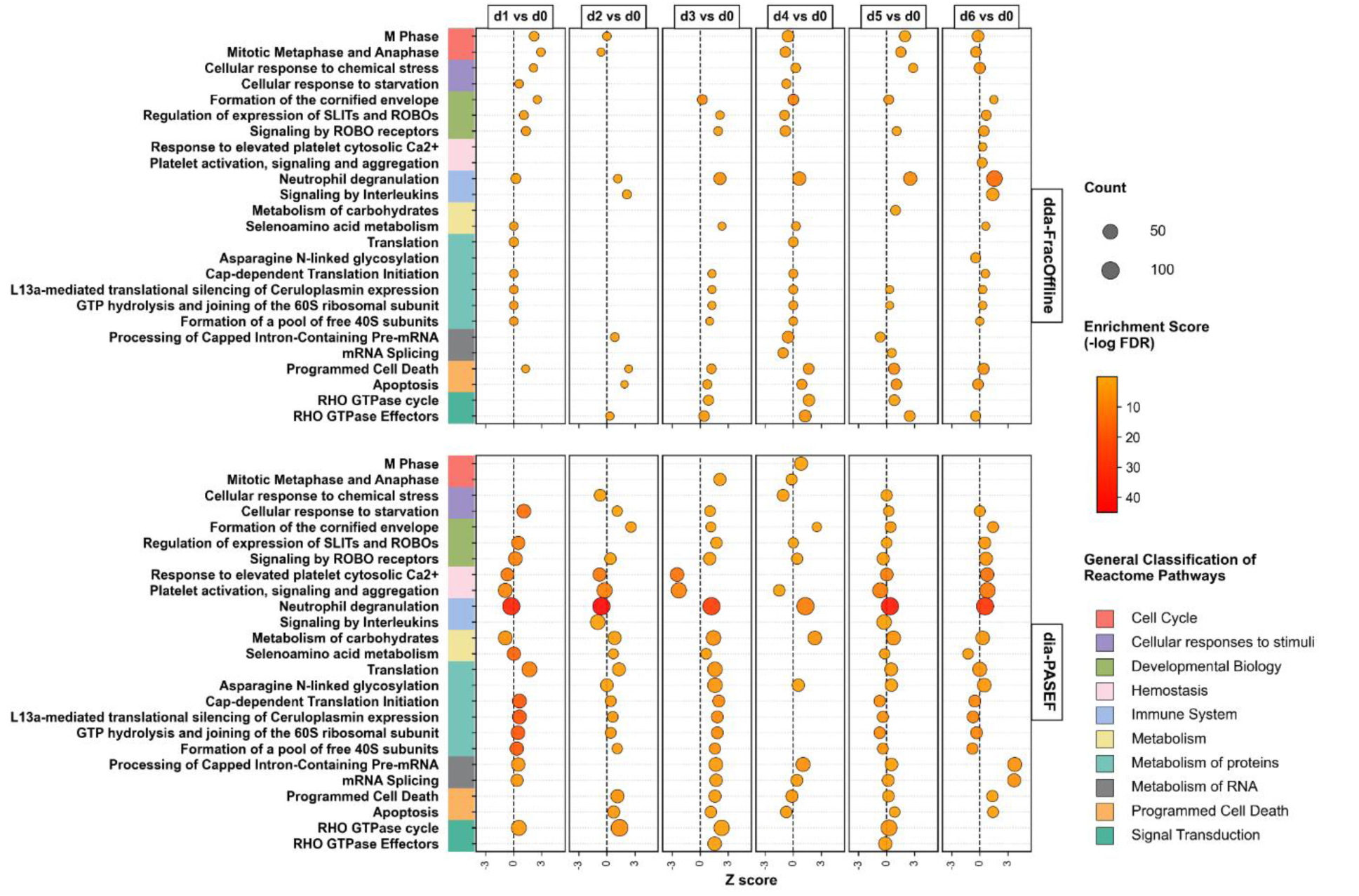
Differentially expressed proteins and biological pathways identified via longitudinal analysis using dda-FracOffline and dia-PASEF. Reactome enrichment analysis of DEPs identified in longitudinal comparisons for each workflow. The Y-axis shows Reactome pathways, and the X-axis represents the directional Z score. Bubble size indicates protein count, and colour indicates the enrichment score (−log FDR).

### 4. Identification of physiological and pathological biomarkers

We evaluated the ability of each strategy to identify known biomarkers of physiological and pathological processes and quantify their abundance. We began with a set of disease biomarkers proposed by the U.S. Food and Drug Administration (FDA-proposed biomarkers) ^42^, focusing on fluids related to various pathologies and examining literature-curated sets of proteins related to female reproductive-tract pathophysiology (Table S1).

#### 4.1 Comparison of FDA-proposed biomarkers via dda-FracOffline and dia-PASEF

The FDA-proposed biomarker panel comprises 121 proteins detectable in biological fluids and associated with female reproductive pathologies ^42^. The ability of each strategy to detect these biomarkers in CVF is compared in Figure 4; 92 and 77 of the 121 proteins, corresponding to 76% and 64%, were quantified by dia-PASEF and dda-FracOffline, respectively; 76 proteins (63%) were quantified by both strategies (Figure 4A). A protein rank-abundance plot demonstrates that dia-PASEF captured proteins across a broader dynamic range than dda-FracOffline (Figure 4B). Sixteen FDA biomarkers were uniquely quantified by dia-PASEF (NT5E, ACE, GLA, C4A, C1QC, C1R, ACP5, PROS1, BRAF, VWF, GALT, MSH2, PKLR, TFRC, TOP2A, and BTD), whereas the protein GFAP was quantified only by dda-FracOffline (Figure 4A). Overall, dia-PASEF showed better quantification consistency and reproducibility than dda-FracOffline for the detection of clinically relevant biomarker candidates in CVF.

**Figure 4.**
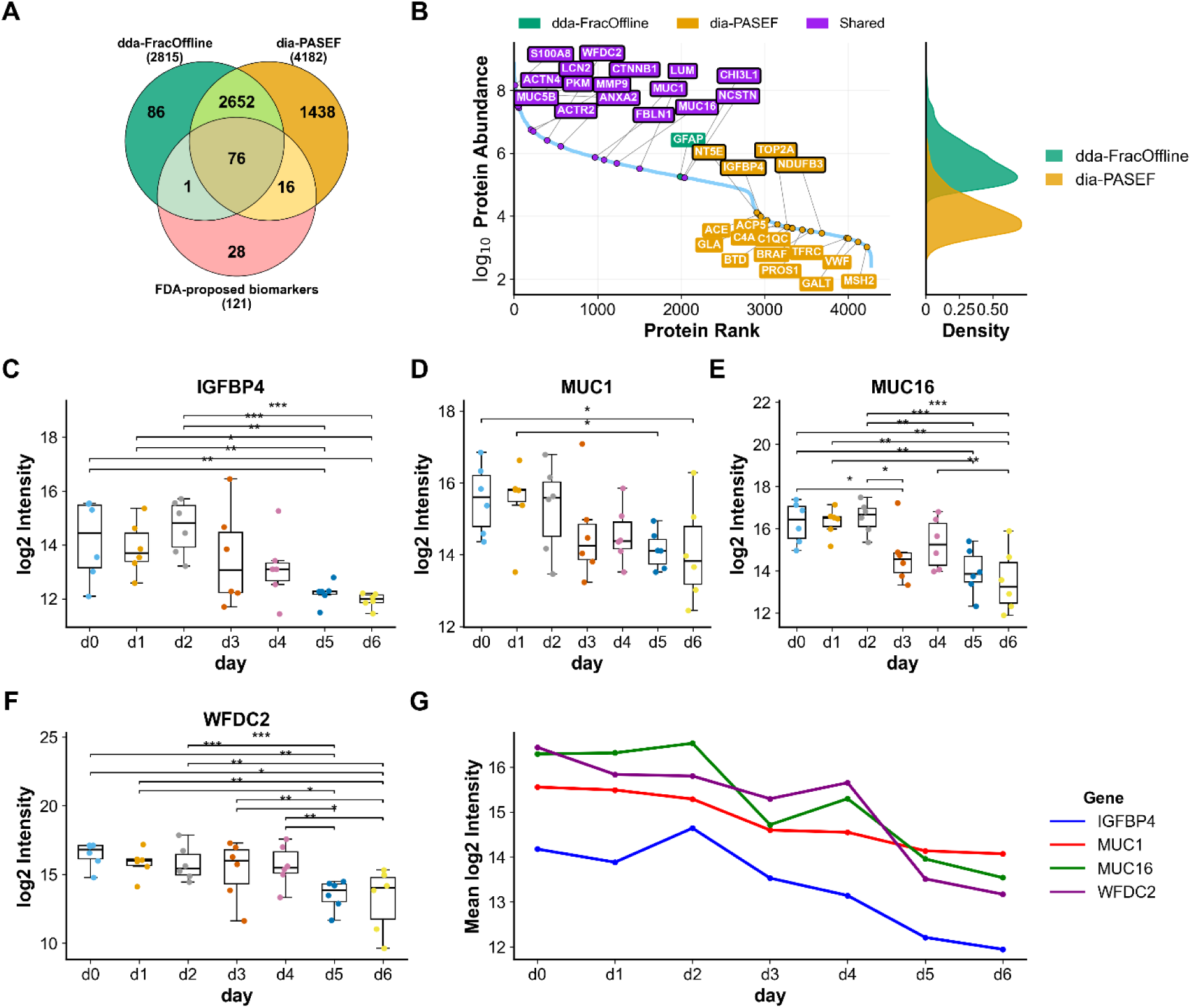
Biomarker identification and longitudinal protein analysis from *d0* to *d6* by dda-FracOffline and dia-PASEF. (A) Venn diagram showing the overlap of quantifiable proteins and FDA-proposed biomarkers, including 76 shared and workflow-specific proteins. (B) Protein abundance rank plot for dda-FracOffline (green), dia-PASEF (orange), and shared proteins (purple). FDA-proposed biomarkers are highlighted, with black-bordered labels indicating proteins included in Table 2; the right panel shows the abundance density distribution. (C, D) Longitudinal abundance of IGFBP4 and MUC1. (E, F) Longitudinal abundance of MUC16 and WFDC2 (***, p < 0.001; **, p < 0.01; *, p < 0.05). (G) Mean longitudinal trajectories of IGFBP4, MUC1, MUC16, and WFDC2.

**Table 2:** Summary of biomarkers associated with female pathologies.

| Pathology | Gene Symbol | Reference(s) |
| --- | --- | --- |
| **Pregnancy pathologies** | *FBLN1* | 43,44 |
|  | *NCSTN* | 45,46 |
|  | *ACTR2* | 45,46 |
|  | *NDUFB3* | 47 |
|  | *IGFBP4* | 48–50 |
| **Cervical carcinoma** | *MUC5B* | 51–53 |
|  | *ANXA2* | 51–54 |
|  | *ACTN4* | 51–54 |
|  | *PKM* | 53,54 |
| **Ovarian carcinoma** | *MUC16* | 55–59 |
|  | *NT5E* | 60 |
|  | *WFDC2* | 9,58–60 |
|  | *KLK5* | 61 |
|  | *MUC1* | 62,63 |
|  | *CHI3L1* | 56,57 |
| **Endometrial carcinoma** | *MUC16* | 64–66 |
|  | *CTNNB1* | 6,64,67,68 |
|  | *WFDC2* | 64,65,68 |
|  | *MMP9* | 64,68,69 |
| **Endometriosis** | *TOP2A* | 70,71 |
|  | *S100A8* | 15,72–74 |
|  | *LCN2* | 15 |
|  | *LUM* | 72,74–76 |

#### 4.2 Selected biomarkers from literature review

A curated list of biomarkers for pregnancy, cervical carcinomas, ovarian carcinomas, endometrial carcinomas, and endometriosis previously reported in the literature is provided in Table 2. Proteins from this curated list are outlined in black in Figure 4B. We examined time-dependent changes in the expression of four representative biomarkers from the curated list to illustrate the longitudinal detection capability of the dia-PASEF workflow (Figure 4C–F; trajectories in Figure 4G). To illustrate the longitudinal capability of dia-PASEF in detecting clinically relevant proteins over repeated measures, we tracked the expression of insulin-like growth factor-binding protein 4 (IGFBP-4), which is implicated in early pregnancy and foetal growth restriction; MUC1, a mucin contributing to reproductive tract integrity; and the following proteins previously investigated as biomarkers of ovarian cancer: CA-125 (MUC16) and HE4 (WFDC2). Insulin-like growth factor-binding protein 4 (IGFBP4) is associated with early pregnancy and foetal growth restriction (Figure 4C, Figure S5). MUC1, expressed throughout the female reproductive tract—including the endocervical, ectocervical, and vaginal epithelia—plays a role in maintaining epithelial barrier function (Figure 4D, Figure S5). Both proteins are physiologically expressed across the menstrual cycle. In addition, the clinically relevant biomarkers CA-125 (Figure 4E, Figure S5) and HE4 (Figure 4F, Figure S5), which are encoded by MUC16 and WFDC2, respectively, were consistently quantified. Longitudinal analysis across the ovulation-anchored repeated-measures window (*d0–d6*) revealed significant day-to-day variation in the expression of all four proteins (Figure 4C–G; Figure S5), supporting the suitability of dia-PASEF for time-dependent biomarker profiling.

## Discussion

CVF is well established as a valuable biological source for research on the pathophysiology of the female reproductive tract. This biofluid contributes to multiple biological processes, including barrier function, immune responses, microbiota preservation, sperm transport, and menstrual cycle regulation. The emergence of advanced proteomics workflows and MS instrumentation provides opportunities for the large-scale identification of reproductive health biomarkers in CVF^5^. In the present study, the performance of dia-PASEF as a proteomic workflow for analysing CVF in longitudinal repeated-measures designs with time-series sampling was evaluated. To our knowledge, this is among the first applications of dia-PASEF for analysing CVF collected across consecutive days of the menstrual cycle, and the results revealed the improved performance of dia-PASEF relative to dda-FracOffline. Comprehensive comparative analyses revealed that dia-PASEF identified more quantifiable proteins with lower across-sample quantitative variability and greater sensitivity for low-abundance proteins. The reduced variability (gCV = 55.6% for dia-PASEF versus 203.1% for dda-FracOffline) and lower proportion of missing values enabled more consistent quantification across samples than dda-FracOffline, improving the robustness of the longitudinal analyses (Figure 2). This interpretation was further supported by the dataset-level analysis, in which total signal intensity showed a stronger association with the major source of variation for dda-FracOffline than for dia-PASEF. These findings indicate that dia-PASEF supports the reliable tracking of protein abundance over time, facilitating the investigation of time-dependent molecular changes in CVF.

Studies of longitudinal effects are essential for assessing the development of physiological phenomena over time, such as in the earliest stages of pregnancy, which remain poorly understood. These analyses are also relevant for monitoring disease progression and therapeutic responses from a pathophysiological perspective. However, longitudinal analysis of CVF remains technically challenging because of the complexity of this biofluid and the limited availability of well-characterised repeated-measures datasets spanning the menstrual cycle. Thus, the application of advanced acquisition strategies such as dia-PASEF represents a methodological advancement, as it provides analytical features suited for complex biofluids and improved detection of low-abundance proteins in time-dependent study designs.

The LC–MS/MS acquisition time was shorter for dia-PASEF, for which we used a single 60-min gradient per sample, than for dda-FracOffline, for which we used a 120-min gradient per fraction. Considering the eight final concatenated fractions, this corresponded to an LC– MS/MS acquisition time of approximately 1 h per sample for dia-PASEF versus 16 h per sample for dda-FracOffline. In addition, the database search time for dia-PASEF was shorter (a few minutes per sample) than that for dda-FracOffline (1–2 hours per sample). These results support the suitability of dia-PASEF for longitudinal repeated-measures proteomic studies.

The higher reproducibility observed for dia-PASEF is consistent with its systematic precursor selection strategy, which enhances proteome depth and sensitivity ^77^. In contrast, the sequential precursor selection inherent to dda-FracOffline is associated with greater variability and reduced efficiency in protein sampling, limiting its applicability in large-scale and time-dependent study designs.

The ability of dia-PASEF to capture comprehensive fragmentation data within a single acquisition cycle contributes to improved protein identification, reduced missing values, and broader proteome coverage. The greater quantitative stability observed with dia-PASEF further supports its application in large-scale longitudinal proteomics, in line with previous reports describing the analytical advantages of data-independent acquisition strategies ^26^.

Although both methodologies allow in-depth protein identification, we observed that dia-PASEF enabled more consistent protein quantification than dda-FracOffline, primarily because the latter showed lower quantifiable protein coverage and a higher proportion of missing values^29^. When dia-PASEF was compared with other DIA approaches applied to CVF, such as SWATH-MS–based methods ^7^, a greater number of proteins was identified and quantified. Previous proteomic studies using SWATH-MS for analysing CVF have reported limitations, including reduced sensitivity for low-abundance proteins, peptide cofragmentation effects ^26^, and higher proportions of missing values.

Enrichment analyses enabled exploration of the functional relevance of the quantified proteins contributing to CVF pathophysiology (Figure 3). Both strategies highlighted pathways related to cellular signalling, protein metabolism, immune responses, interleukin signalling, mitotic metaphase–anaphase transition, mRNA splicing, and RHO GTPase signalling, which participate in reproductive system biology ^78,79^. dia-PASEF showed higher enrichment scores across the time-dependent window, suggesting improved biological coverage. For example, neutrophil gelatinase-associated lipocalin (NGAL; encoded by LCN2), which is involved in neutrophil degranulation ^15^ and signalling pathways, was consistently quantified (Figure 3, Figure 4B).

Tissue enrichment analysis confirmed the biological relevance of the quantified proteins, highlighting their associations with tissues of the female reproductive system ^80^, including the uterus and uterine cervix. These findings are consistent with the ability of dia-PASEF to provide system-specific sensitivity and enable time-dependent biological profiling, particularly for reproductive and immune-related functions in longitudinal CVF proteomic analyses (Figure S4).

We compared proteins quantified by dia-PASEF and dda-FracOffline with those in the FDA-proposed biomarker panel ^42^. Among the FDA-proposed biomarkers, 76 proteins were quantified by both workflows; in addition, dia-PASEF quantified 16 unique proteins, whereas dda-FracOffline quantified one unique protein (Figure 4A). The dia-PASEF workflow quantified known biomarkers across a broader dynamic range, including both high- and low-abundance proteins (Figure 4B), supporting its suitability for comprehensive biomarker profiling of CVF. These findings indicate improved detection performance of dia-PASEF for FDA-proposed biomarkers within this longitudinal repeated-measures design.

Low-abundance proteins were linked to biological processes relevant to menstrual cycle regulation and gynaecological diseases. For example, C1QC and VWF are involved in immune responses and vascular regulation. A broader and more uniform dynamic range was observed for dia-PASEF, reflecting improved signal-to-noise performance, a feature that is advantageous for longitudinal analyses of complex biofluids^34^ such as CVF. As an additional strategy for evaluating clinically relevant biomarkers, dia-PASEF facilitated the detection of low-abundance proteins across the time-dependent repeated-measures window. IGFBP-4 (Figure 4C), whose levels have been associated with early pregnancy and foetal growth restriction ^48,50^, and MUC1 (Figure 4D), a mucin expressed throughout the female reproductive tract— including the endocervical, ectocervical, and vaginal epithelia—were consistently quantified. MUC1 plays a role in maintaining epithelial integrity and barrier function ^62,63^.

We also quantified proteins associated with pathological processes, including alpha-actinin-4 (ACTN4) and annexin A2 (ANXA2), in CVF, which have been reported in previous studies ^51–53^ (Table 2). Two proteins clinically relevant to gynaecological carcinomas were quantified: cancer antigen 125 (CA-125), which is encoded by the MUC16 gene (Figure 4E), and human epididymis protein 4 (HE4), which is encoded by the WFDC2 gene (Figure 4F). These proteins were quantified in CVF samples collected across the ovulation-anchored window (*d0–d6*). Both biomarkers have been widely investigated in ovarian^58,59^ and endometrial carcinoma^64,65^. Longitudinal analysis revealed significant day-to-day variation in the expression of these proteins across the repeated-measures window (Figure S5). These findings suggest that dia-PASEF facilitates time-dependent monitoring of the abundance of clinically relevant proteins in CVF, supporting its potential application in studies of pathologies related to the female reproductive tract.

Proteins present in CVF may also represent promising candidates for longitudinal pathophysiological markers. Because dia-PASEF yielded a reduced proportion of missing values and enabled more consistent time-dependent protein profiling, it generated a more complete quantitative dataset (Figure 2A). These characteristics support its application for tracking protein dynamics during physiological processes, such as endometrial receptivity, as well as in pathological contexts, over time. A consistent pattern of protein variation was observed across specific days of the cycle (Figure 4G, Figure S5), providing insight into defined physiological windows within the ovulation-anchored repeated-measures framework. These findings support the longitudinal monitoring of protein abundance in CVF under physiological conditions and suggest potential future applications of dia-PASEF in disease or therapeutic monitoring, which would require validation in appropriate clinical cohorts. No pooled QC or reference samples were included; therefore, instrument performance and batch effects could not be independently assessed. The stronger association with total signal intensity observed for dda-FracOffline may reflect its greater analytical complexity but cannot be attributed specifically to instrument batch effects.

In conclusion, this study provides evidence supporting the suitability of dia-PASEF as an analytical platform for longitudinal repeated-measures proteomic analysis of CVF, addressing limitations inherent to DDA-based workflows and supporting future research on biomarker discovery for fertility, early pregnancy, and gynaecological health and disease. Overall, this approach provides a scalable methodological framework for time-dependent proteomic analyses of complex reproductive biofluids.

## Limitations

The principal limitation of this study is the small number of participants, which resulted from the stringent selection criteria applied when selecting individuals from the preexisting EARLY-PREG biobank. However, the study was designed as a methodological validation of the dia-PASEF workflow for longitudinal proteomic analysis of CVF rather than as a population-based biological study. Consequently, the primary objective was to evaluate the analytical performance, reproducibility, proteome coverage of the workflow, and its ability to detect temporal proteomic changes under highly controlled conditions.

Although only six women were included, due to the repeated-measures design, 42 longitudinal CVF samples were collected, with each participant serving as her own control. This design minimised between-subject variability and provided an appropriate framework for comparing the analytical performance of the proteomic workflows. Accordingly, the differentially expressed protein and pathway analyses should be regarded as exploratory and should not be interpreted as population-level biological findings. Larger prospective studies are needed to validate these observations in more diverse cohorts. Nevertheless, this work shows the suitability of dia-PASEF as a robust platform for longitudinal repeated-measures proteomic analysis of CVF and establishes a methodological framework for future biomarker discovery studies in the field of reproductive health.

## Data Availability

All data produced in the present study are available upon reasonable request to the authors

## Authors’ roles

E.S.K. led the study. M.H. and E.S.K. drafted the manuscript and conceptualised the study. E.S.K., M.H., and C.V. contributed to the study design. M.H. designed, supervised, and performed the proteomics methodology. P.S., B.A., and V.L. contributed to study conception. M.H., P.S., and G.N. developed the experimental system, performed the experiments, and analysed the data. F.C. and G.N. supported the experimental work. M.H., P.S., B.A., and V.L. reviewed and commented on the drafts of the manuscript. E.S.K. revised the manuscript and approved the final version. E.S.K. is the guarantor of the study and accepts full responsibility for the integrity of the work.

## Acknowledgements

The authors thank Dr. Ute Woehlbier (Center for Integrative Biology, Universidad Mayor) for English language editing. We also acknowledge Estefanía Nova-Lamperti (Molecular and Translational Immunology Laboratory, Department of Clinical Biochemistry and Immunology, Faculty of Pharmacy, Universidad de Concepción, Chile) for assisting with figure design. I.F.-M. received professional service fees from Fundación de Investigación San Ramón (FISAR), Concepción, Chile.

## Funding

This work is part of the EARLY-PREG preconception open cohort study and was supported by research grants awarded by Fundación de Investigación San Ramón (FISAR), based in Chile. The pilot study and first recruitment wave were supported by grants #MEL109112011 and #MEL109112011R4 awarded to E.S.K., C.V., and J.F.S. The second recruitment wave was supported by grants #MEL109112011R5 and #MEL131032017R1 awarded to E.S.K. The third wave was supported by grant #MEL205062018 awarded to E.S.K. and M.H. Current funding for the fourth recruitment wave and mass spectrometry research on maternal CVF is supported by grant REH042024-01 awarded to M.H., G.N., and E.S.K.

## Conflict of interest

E.S.K. serves in an unpaid honorary capacity as a research advisor and reviewer for Fundación de Investigación San Ramón (FISAR). No other competing interests are declared.

## Notes

### Competing Interest Statement

E.S.K. serves in an unpaid honorary capacity as a research advisor and reviewer for Fundacion de Investigacion San Ramon (FISAR). No other competing interests are declared.

### Author Declarations

The ethics committee of the Servicio de Salud Concepcion, Biobio region, Chile gave ethical approval for this work (CODE: 17-03-06 / 26-01-04).

### Summary of Updates

The wording was improved, and responses were provided to questions raised by reviewers.

